# Risk of fomite-mediated transmission of SARS-CoV-2 in child daycares, schools, and offices: a modeling study

**DOI:** 10.1101/2020.08.10.20171629

**Authors:** Alicia N.M. Kraay, Michael A. L. Hayashi, David M. Berendes, Julia S. Sobolik, Juan S. Leon, Benjamin A. Lopman

## Abstract

SARS-CoV-2 can persist on surfaces, suggesting that surface-based transmission might be important for this pathogen. We find that fomites may be a substantial source of risk, particularly in schools and child daycares. Combining surface cleaning and decontamination with strategies to reduce pathogen shedding on surfaces can help mitigate this risk.

## Main text

SARS-CoV-2 can be transmitted through direct droplet spray and close contact, but also persists for up to 72 hours on surfaces [1, 2], suggesting that transmission might also occur through contaminated surfaces, hereafter called ‘fomites.’

Conventional epidemiologic studies cannot distinguish between competing transmission pathways when they act simultaneously. Therefore, we use a transmission modeling approach to explore the potential for fomite transmission, without other pathways. We adapted a published fomite transmission model [3] for SARS-CoV-2. In our model, individuals are classified as susceptible, infectious, or recovered. Infectious individuals shed pathogens onto fomites or hands, but only a fraction of surfaces (λ) are accessible for contamination. Hands may become contaminated from viral excretion or from touching virally-contaminated fomites. Susceptible individuals may become infected through touching their face and mouth with contaminated hands (see Appendix).

Using this model, we explore how fomite transmission varies by location (comparing child daycares, schools and offices), disinfection strategy, and surface type. While precise parameter values are likely to vary on a case-by-case basis, child daycares are assumed to have higher frequency of fomite touching and the fraction of surfaces susceptible to contamination than offices, whereas schools are intermediate for both factors [3].

We considered the following surface cleaning and disinfection frequencies: every 8 hours (once/workday), every 4 hours (twice/workday), and hourly. We also considered handwashing interventions, but they had minimal impact and are not included in our main results (see Appendix). Because SARS-CoV-2 persistence varies by surface, we compared transmission for stainless steel, plastic, and cloth. For unknown persistence and transfer efficiency parameters, we used influenza values because SARS-CoV-2, like influenza, is an enveloped virus, which tend to have lower persistence than non-enveloped viruses on surfaces [4]. For uncertain infectivity parameters, we used rhinovirus values, because of the similarity in symptoms likely to drive transmission and because of the similarity in dose-response curves between coronaviruses and rhinoviruses [5, 6]. Early studies on shedding rates for infectious virus had small sample sizes and thus uncertain estimates [7]. Additionally, shedding rates are likely to vary based on mask wearing practices [8]. We varied shedding rate estimates in sensitivity analyses to capture variability due to both factors (see Appendix). In our model, situations where the basic reproduction number (R_0_) for the fomite route exceeds 1 could sustain ongoing transmission in a given setting, whereas transmission could be interrupted when R_0_ falls below 1. We explored what interventions could interrupt fomite transmission.

Our estimates suggest that fomite transmission could sustain SARS-CoV-2 transmission in many settings. The fomite R_0_ ranged from 2 in low risk venues (offices) to about 20 in high risk settings such as child daycares (Figure 1A). SARS-CoV-2 transmission risk is higher than both influenza and rhinovirus. However, rhinovirus has slightly higher transmission than SARS-CoV-2 in some venues when touching rates are low because of its higher persistence on hands and transfer efficiencies.

**Figure 1A.**
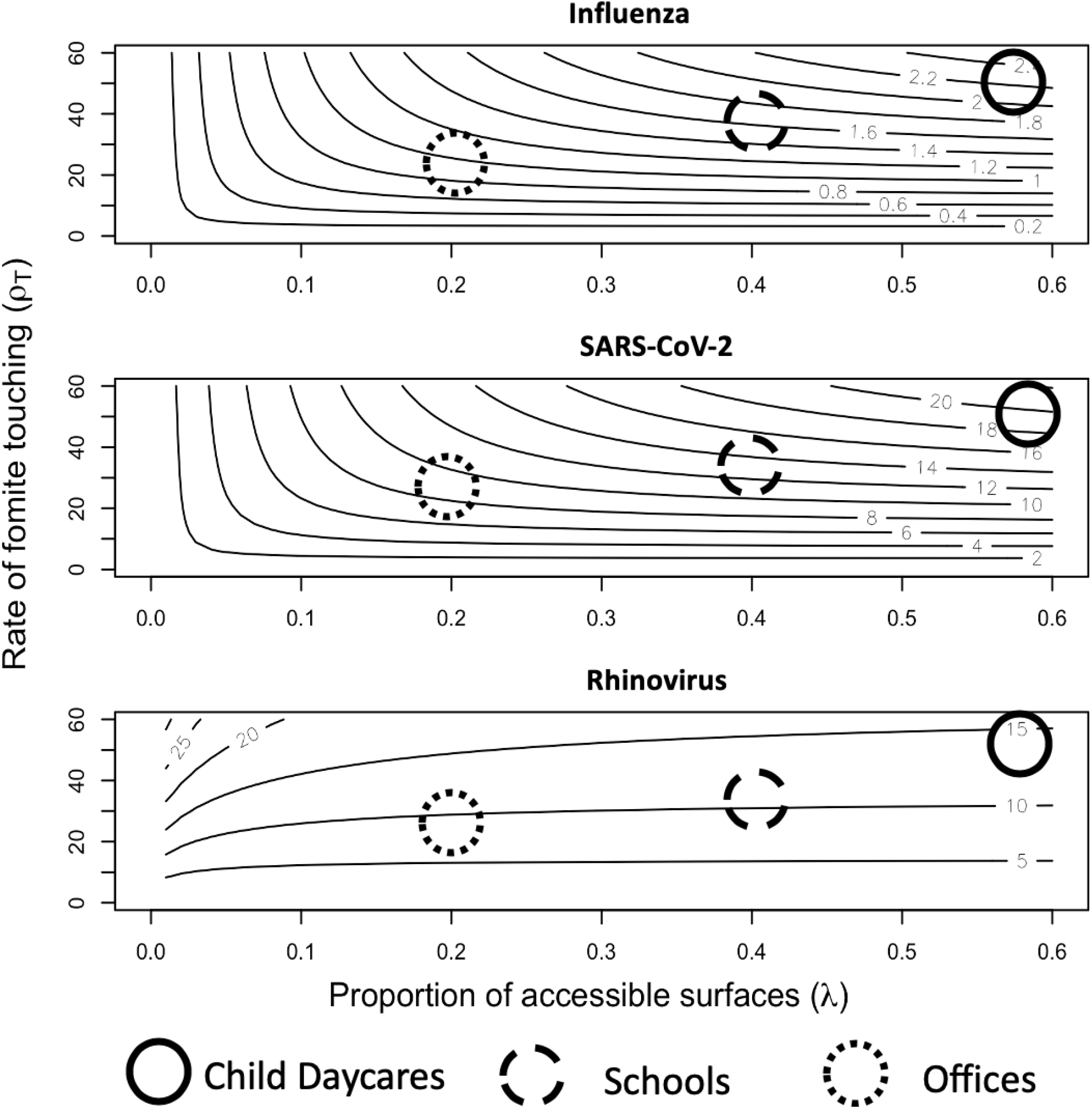
Predicted basic reproduction number for the fomite pathway without any interventions for child daycares, schools, and office locations. Touching rates and accessible surfaces are not known precisely, so larger circular symbols are used to reflect uncertainty, highlighting the plausible range. All three pathogens are shown using decay rates from stainless steel surfaces. Parameters used for each pathogen are shown in Table S1.

We found that hourly cleaning and disinfection alone could bring the fomite R_0_ below 1 in some office settings, particularly combined with reduced shedding, but would be inadequate in child daycares and schools (Figure 1B, Figure S3). If viral shedding can be reduced, surface cleaning and decontamination would be effective in a wide range of settings, but child daycares would remain at high risk. Decay rates on cloth were high and are unlikely to sustain transmission. Therefore, cleaning and disinfection frequencies could be differential by surface, with hourly interventions being helpful for frequently touched non-porous surfaces, with porous surfaces (like plush toys) being cleaned and disinfected less frequently. In child daycares, intervening directly after high risk shedding events (e.g., a feverish individual coughs directly on a surface) in addition to intervening at standard intervals (such as hourly) might be beneficial.

**Figure 1B.**
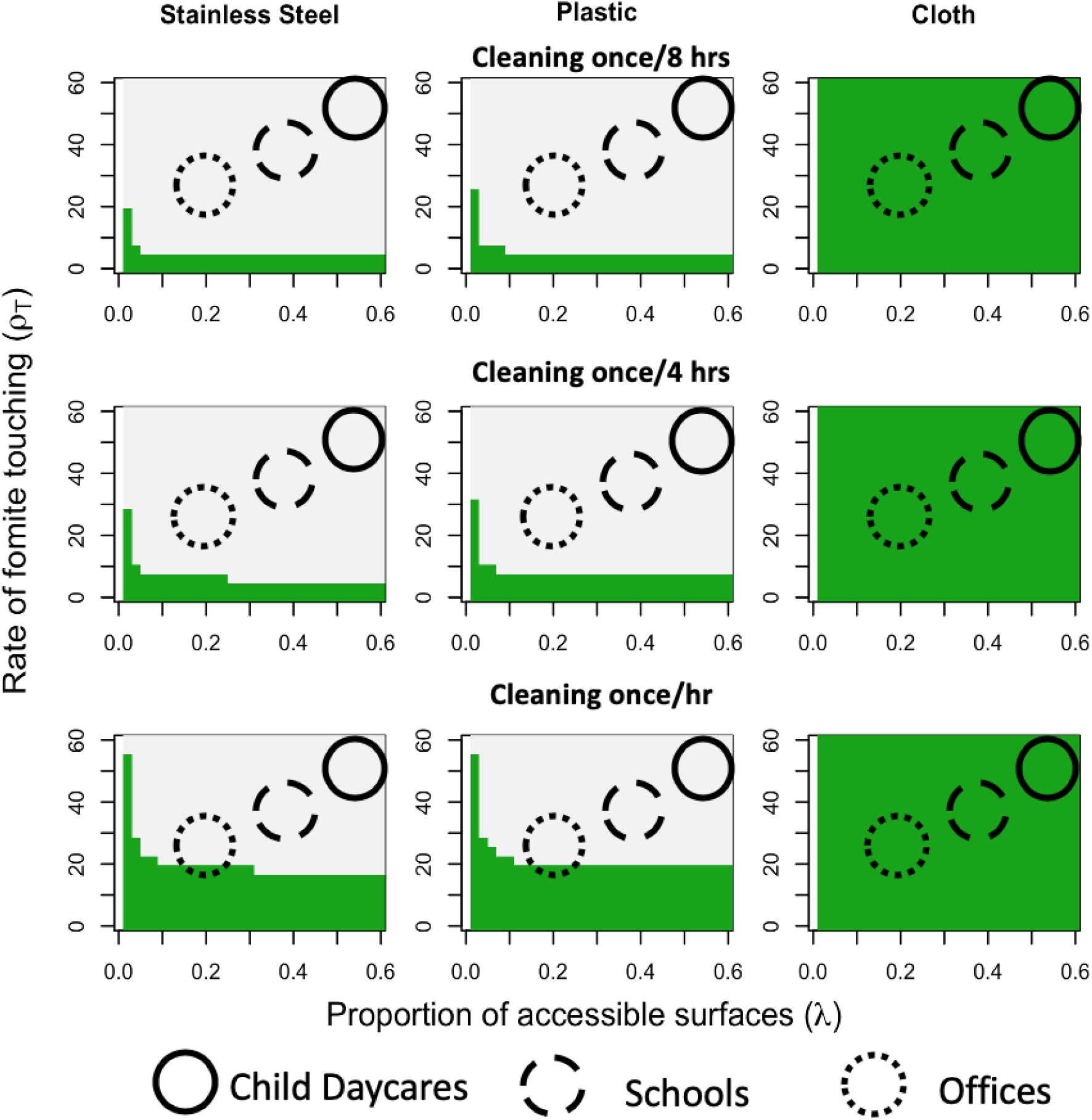
Reductions in the basic reproduction number for different surfaces. For areas in green, the projected reproduction number from fomite transmission is below 1. For comparison, cleaning every 2 hours was considered as a sensitivity analysis (see Appendix).

These results assume that outbreaks begin with a single infectious case as opposed to many cases being introduced simultaneously. Thus, these results apply when SARS-CoV-2 incidence is low, which might be achievable in individual locations even if community incidence is high. Near the epidemic peak, more detailed simulations are needed because environmental contamination would likely exceed the linear range of the dose-response curve [9], leading to an overestimation of the risk of fomite transmission. Because our objective was to assess the potential impact of fomite-mediated transmission alone, we did not account for direct transmission through direct droplet spray, aerosols, or hand-to-hand contact, all of which are likely major contributors to transmission in many settings [10]. While direct transmission is important, our model suggests fomites can also transmit, which is important for exposures that are not in-person. For simplicity, we do not model asymptomatic and symptomatic infections separately, assuming that fomite transmission is similar for both groups (see Appendix).

Overall, fomite transmission may be an important source of risk for SARS-CoV-2. However, frequent cleaning and disinfection can reduce this risk.

## Data Availability

All data used in the manuscript are taken from published studies cited in the text.

## Disclaimer

The findings and conclusions in this report are those of the authors and do not necessarily represent the official position of the Centers for Disease Control and Prevention.

